# Pharmacometabolomics in TB Meningitis – understanding the pharmacokinetic, metabolic, and immune factors associated with anti-TB drug concentrations in cerebrospinal fluid

**DOI:** 10.1101/2023.12.14.23299982

**Authors:** Jeffrey M. Collins, Maia Kipiani, Yutong Jin, Ashish A. Sharma, Jeffrey A. Tomalka, Teona Avaliani, Mariam Gujabidze, Tinatin Bakuradze, Shorena Sabanadze, Zaza Avaliani, Henry M. Blumberg, David Benkeser, Dean P. Jones, Charles Peloquin, Russell R. Kempker

## Abstract

Poor penetration of many anti-tuberculosis (TB) antibiotics into the central nervous system (CNS) is thought to be a major driver of morbidity and mortality in TB meningitis (TBM). While the amount of a particular drug that crosses into the cerebrospinal fluid (CSF) varies from person to person, little is known about the host factors associated with interindividual differences in CSF concentrations of anti-TB drugs. In patients diagnosed with TBM from the country of Georgia (n=17), we investigate the association between CSF concentrations of anti-TB antibiotics and multiple host factors including serum drug concentrations and CSF concentrations of metabolites and cytokines. We found >2-fold differences in CSF concentrations of anti-TB antibiotics from person to person for all drugs tested including cycloserine, ethambutol, imipenem, isoniazid, levofloxacin, linezolid, moxifloxacin pyrazinamide, and rifampin. While serum drug concentrations explained over 40% of the variation in CSF drug concentrations for cycloserine, isoniazid, linezolid, and pyrazinamide (adjusted R^2^>0.4, p<0.001 for all), there was no evidence of an association between serum concentrations of imipenem and ethambutol and their respective CSF concentrations. CSF concentrations of carnitines were significantly associated with concentrations of ethambutol and imipenem (q<0.05), and imipenem was the only antibiotic significantly associated with CSF cytokine concentrations. These results indicate that there is high interindividual variability in CSF drug concentrations in patients treated for TBM, which is only partially explained by differences in serum drug concentrations and not associated with concentrations of cytokines and chemokines in the CSF.

## Introduction

Tuberculosis meningitis (TBM) is the most lethal manifestation of tuberculosis (TB) disease, with a mortality rate ≥ 25% in those with drug susceptible disease and >65% in persons with drug resistance.^1,2^ Therapeutic options for TBM are more limited than in other forms of TB due to poor penetration of many anti-TB drugs into the central nervous system (CNS).^1,3–5^ However, the concentration of antibiotics in the CNS can vary widely by antibiotic and from person to person, indicating certain endotypes exist that lead to favorable CNS penetration of drugs. While it is generally thought that meningeal inflammation increases the CNS concentration of antibiotics, there is limited empirical evidence supporting this concept in TBM.^6^ Further, little is known about the specific inflammatory signaling networks in TBM that may modulate CNS penetration of anti-TB chemotherapeutic agents. An improved understanding of host responses associated with CNS penetration of anti-TB drugs could help inform new strategies to enhance drug delivery to the site of disease in TBM.

Recent advances in metabolomics and high-density cytokine measurement allow for high-resolution immunometabolic profiling of a variety of human samples including cerebrospinal fluid (CSF).^7^ Such profiling allows for simultaneous measurement of over 30 cytokines and thousands of metabolites, including xenobiotics^8^ and molecules that regulate inflammation.^9,10^ Combining measurement of these host response molecules with drug concentrations at the site of disease has the potential to elucidate metabolic and inflammatory processes that regulate drug concentrations and efficacy in TBM; an approach termed pharmacometabolomics.^11^

In a well characterized population of patients from the country of Georgia diagnosed and treated for TBM, we integrated data on serum and CSF concentrations of anti-TB antibiotics^5^ and CSF concentrations of metabolites and cytokines.^7^ By integrating these data sets, we sought to address multiple knowledge gaps that currently exist with regard to antibiotic therapy for TB meningitis as follows: 1) the degree of interindividual variability in CSF drug concentrations, 2) whether variation in serum drug concentrations explain the variation in CSF drug concentrations, 3) whether untargeted metabolomics can be used to accurately measure concentrations of TB antibiotics in the CNS, and 4) whether individual differences in the CSF milieu of soluble immunometabolic mediators (i.e. metabolites and cytokines) are associated with CSF drug concentrations of different anti-TB drugs.

## RESULTS

### Participants

We studied CSF samples from 17 participants with suspected TBM presenting to the National Center for Tuberculosis and Lung Diseases (NCTLD) (**Table 1**). Among the 17 patients with suspected TBM, five (29%) were microbiologically confirmed while three (18%) were considered to have probable TBM and nine (53%) were considered to have possible TBM based on clinical and laboratory criteria.^12,13^ The median CSF white blood cell (WBC) count at diagnosis was 209 cells/mm^3^ (IQR 130-286), with 94% lymphocytes (IQR 85-96), a median glucose of 40 mg/dL (IQR 21-50), and total protein of 99 mg/dL (IQR 66-132). Antibiotic regimens were selected by treating clinicians based on NCTLD guidelines and response to treatment as previously reported.^5,14^ Five persons with TBM were deemed to have a poor initial response to treatment, two of whom were diagnosed with multi-drug-resistant (MDR)-TBM based on microbiologic testing.

**Table 1:**

|  |  |
| --- | --- |
|  | TB meningitis (n=17) |
| Age, median (IQR) | 36 (31-44) |
| Female sex, n (%) | 9 (53) |
| TB meningitis diagnostic category, n (%) |  |
| Definite (microbiologically confirmed) | 5 (29) |
| Probable | 3 (18) |
| Possible | 9 (53) |
| CSF total WBC, median (IQR) | 209 (130-286) |
| % Lymphocytes, median (IQR) | 94 (85-96) |
| % Neutrophils, median (IQR) | 5 (2-8) |
| TB meningitis grade, n (%) |  |
| Grade 1 | 8 (47) |
| Grade 2 | 9 (53) |
| Grade 3 | 0 (0) |

### Variation in CSF Concentrations of Anti-TB Drugs

We first sought to characterize the interindividual variability of anti-TB drug concentrations in the CSF. As per standard of care for patients hospitalized with TBM at NCTLD, lumbar punctures were performed at diagnosis (baseline) and approximately 7, 14, and 28 days after TBM treatment initiation and monthly for as long as patients were hospitalized to follow CSF cell and protein counts in response to treatment.^5,14^ Due to differences between participants in the number of antibiotic doses received at the time of the baseline CSF sample, only samples obtained ≥7 days after treatment start were included in this analysis. Antibiotic concentrations were quantified in the serum and CSF at each time point either 2 or 6 hours after the most recent antibiotic dose as previously described.^5^ A 2- to 4-fold difference in CSF drug concentrations was found between participants at 2 hours (**Figure 1A-I**) and 6 hours (**Figure 2A-I**) after the dosing of anti-TB antibiotics. This was observed across antibiotics, indicating a large amount of interindividual variability in CSF drug concentrations regardless of the drug used.

**Figure 1 –.**
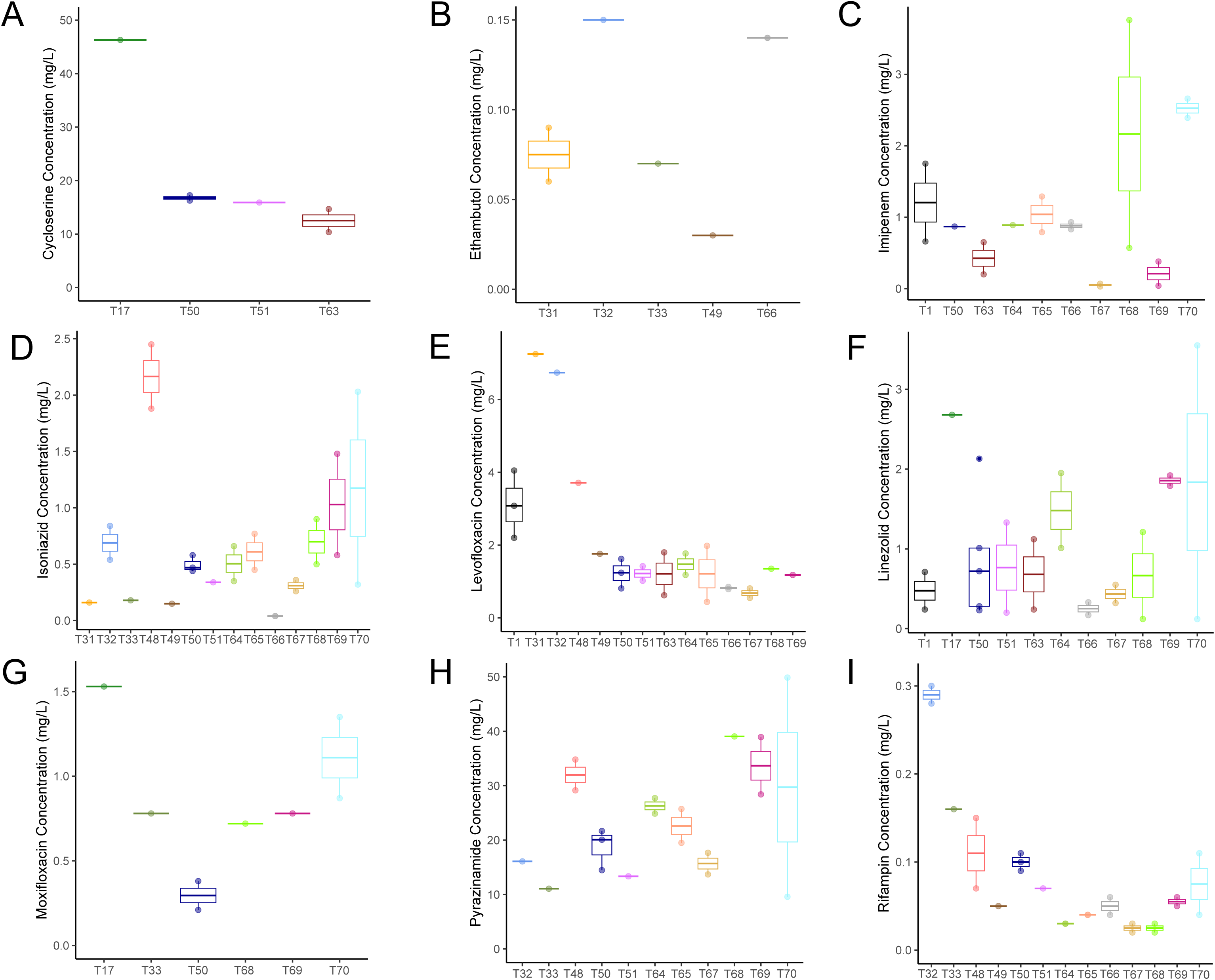
Boxplots depicting median cerebrospinal fluid concentrations of anti-TB antibiotics 2 hours after the most recent antibiotic dose for 17 study participants: (A) cycloserine, (B) ethambutol, (C) imipenem, (D) isoniazid, (E) levofloxacin, (F) linezolid, (G) moxifloxacin, (H) pyrazinamide, and (I) rifampin.

**Figure 2 –.**
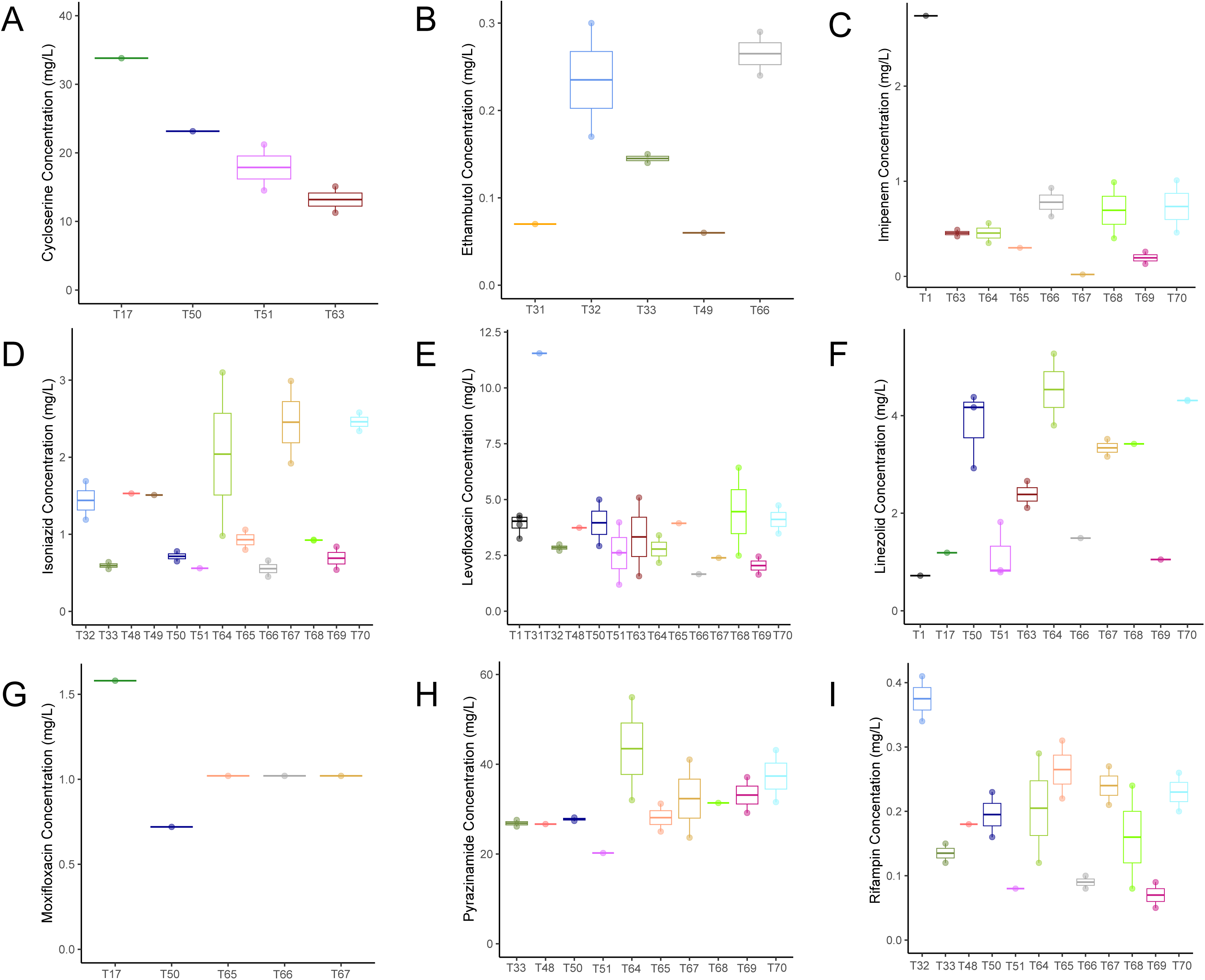
Boxplots depicting median cerebrospinal fluid concentrations of anti-TB antibiotics 6 hours after the most recent antibiotic dose for 17 study participants: (A) cycloserine, (B) ethambutol, (C) imipenem, (D) isoniazid, (E) levofloxacin, (F) linezolid, (G) moxifloxacin, (H) pyrazinamide, and (I) rifampin.

There are multiple sources of potential interindividual variability in CSF drug concentrations among patients with TBM including integrity of the blood-brain and blood-CSF barriers, variability in protein binding, and differential expression of drug transporters.^15^ TB drugs are also known to have large differences in serum concentration from person to person, which could explain some or all of the variability in CSF drug concentrations.^16^ To examine this possibility, we sought to determine how well concomitantly collected serum drug concentrations explained the variation in CSF drug concentrations using a mixed effects linear model. We found the relationship between plasma and CSF drug concentrations varied substantially from drug to drug. For cycloserine, isoniazid, and linezolid, we calculated an adjusted R^2^ of >0.5, suggesting serum drug concentrations explained over 50% of the variability in CSF drug concentrations (**Figure 3A-C**). For other drugs including pyrazinamide, levofloxacin, and rifampin, serum drug concentrations explained a statistically significant amount of variation in CSF concentrations, but the adjusted R^2^ values were 0.4, 0.23 and 0.19 respectively (**Figure 3D-F**). For moxifloxacin, imipenem, and ethambutol, serum drug concentrations were not significantly associated with CSF drug concentrations (**Figure 3G-I**).

**Figure 3 –.**
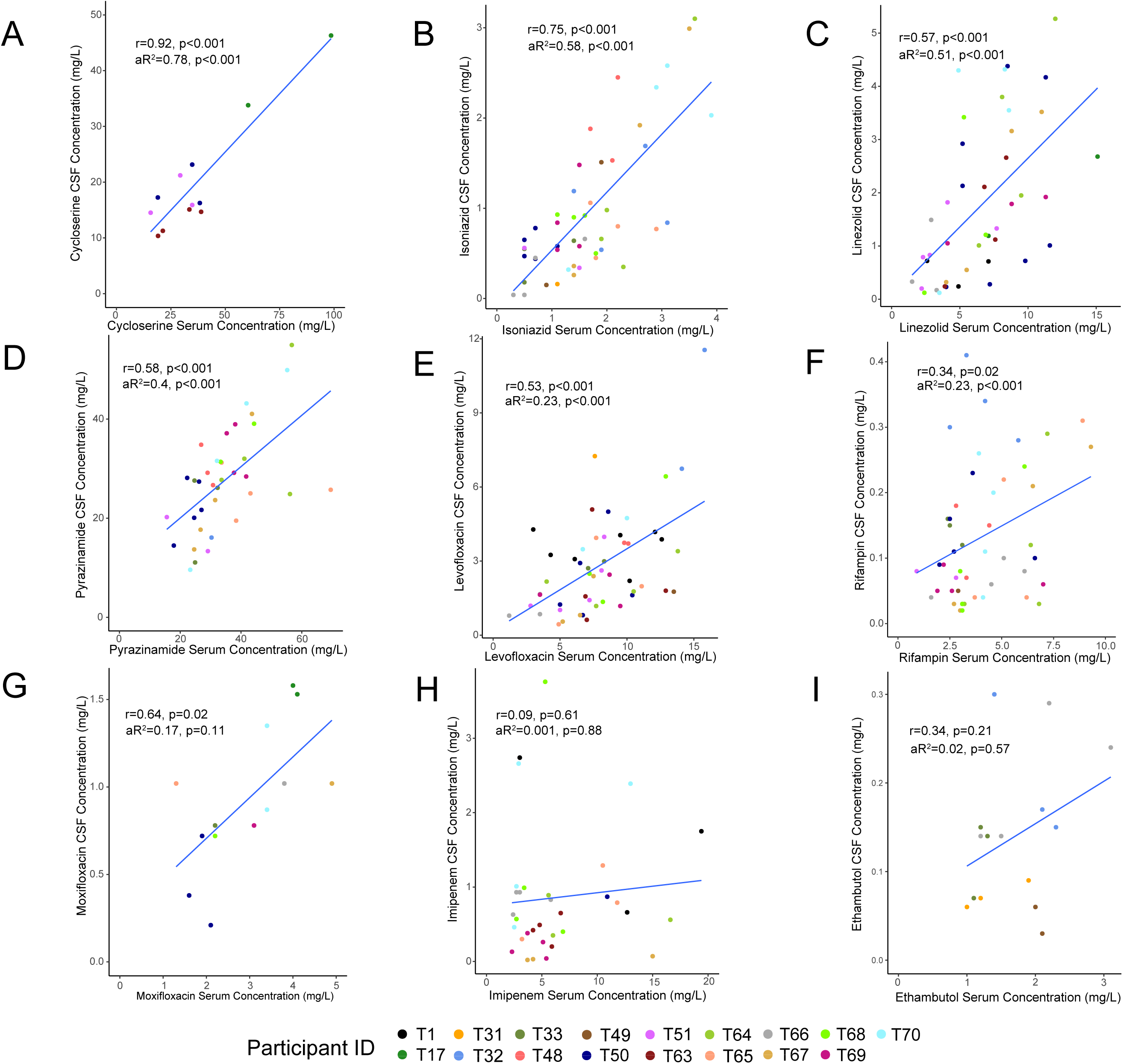
Scatter plots with fitted regression lines depicting the relationship between serum and cerebrospinal fluid concentrations of anti-TB drugs: (A) cycloserine, (B) isoniazid, (C) linezolid, (D) pyrazinamide, (E) levofloxacin, (F) rifampin, (G) moxifloxacin, (H) imipenem, and (I) ethambutol. For each figure, r represents the correlation coefficient with corresponding p-value. The adjusted R^2^ was calculated using a mixed effects linear model, adjusting for sampling time (2 or 6 hours), with p-values derived from the pairwise comparison of models including and excluding the serum concentration of each drug.

### Using Untargeted Metabolomics to measure CSF Drug Concentrations

We performed untargeted high-resolution metabolomics on CSF samples using combined liquid chromatography-mass spectrometry in dual ionization mode with HILIC positive and c18 negative chromatography (see Methods for full details). We detected 6,596 metabolic features in positive ionization mode and 9,427 in negative ionization mode. We then performed a targeted search for molecular features that were within a mass and retention time error range of 5 ppm and 30 seconds respectively for the M+H or M-H adducts of anti-TB drugs. This query yielded high confidence matches for several anti-TB drugs including ethambutol, isoniazid, pyrazinamide, rifampin, linezolid, imipenem, levofloxacin, and moxifloxacin. We then performed a correlation analysis of the peak intensity for each annotated metabolite and the antibiotic concentration as measured using MS/MS and a purified standard curve.^5^ We found that the intensity value for each antibiotic as measured by our untargeted metabolomics platform was strongly correlated with the absolute concentration of most anti-TB antibiotics in the CSF. Peaks for ethambutol, pyrazinamide, isoniazid, linezolid, and imipenem all demonstrated a Pearson correlation coefficient of >0.7 when compared with absolute drug concentrations (**Figure 4A-E**; p<0.001 for all). The peak intensity for moxifloxacin (r=0.53, p=0.05) and levofloxacin (r=0.45, p<0.001), as well as rifampin (r=0.48, p<0.001), were also significantly correlated with measured CSF concentrations, but the relationship was less strong than for the other antibiotics studied (**Figure 4F-H**). In the case of rifampin, this may have been due in part to the relatively low concentration of this antibiotic in the CSF. These data indicate that untargeted metabolomics can be an effective method to approximate many anti-TB drug concentrations in biofluids.

**Figure 4 –.**
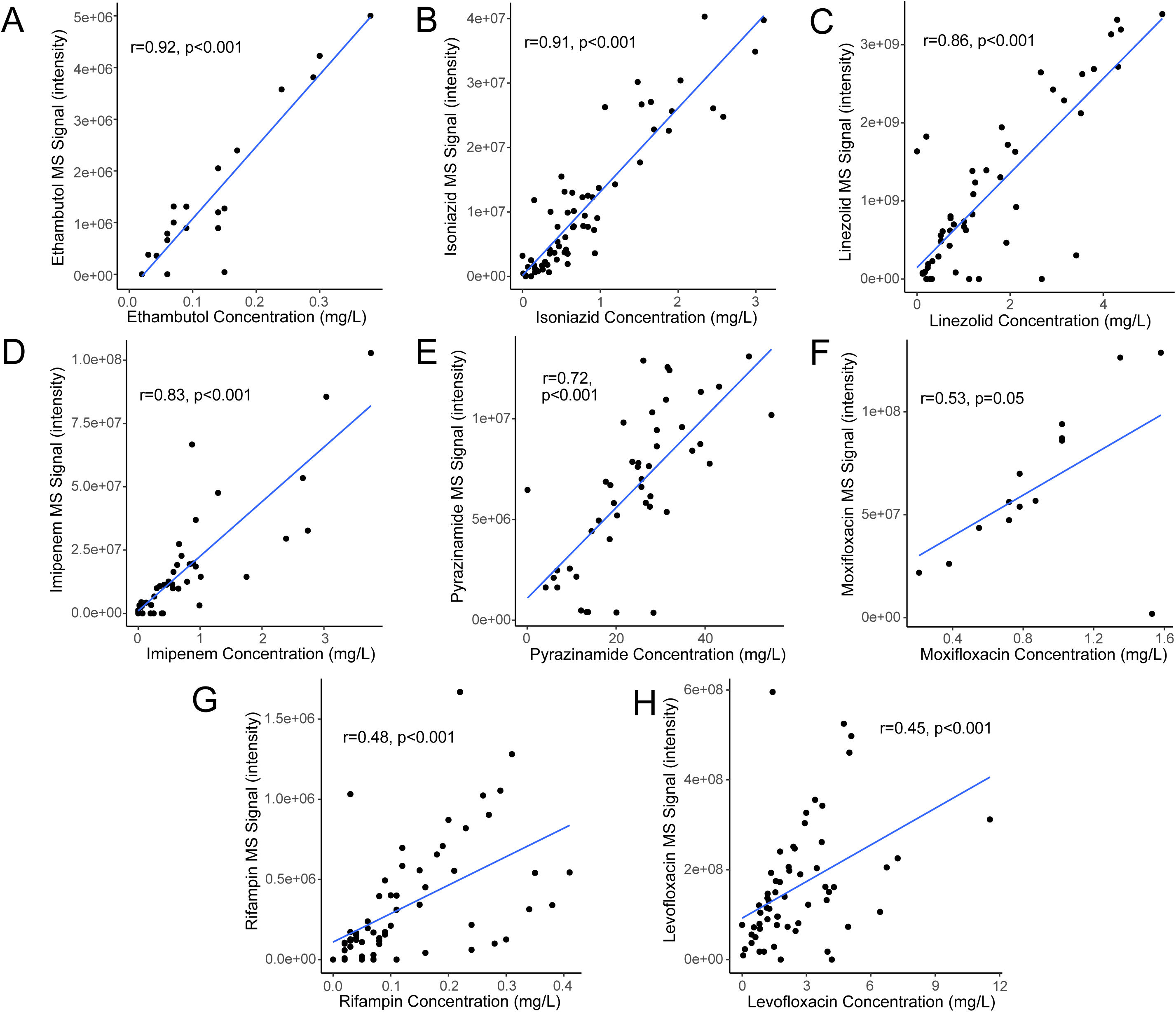
Scatter plots with fitted regression lines depicting the relationship between cerebrospinal fluid concentrations of anti-TB drugs and their mass spectrometry intensity value as measured on the untargeted metabolomics platform: (A) ethambutol, (B) isoniazid, (C) linezolid, (D) imipenem, (E) pyrazinamide, (F) moxifloxicin, (G) rifampin, and (H) levofloxacin. For each figure, r and p values are based on the Pearson correlation coefficient for each drug.

### Relationship Between CSF Metabolism, Inflammation, and Drug Concentrations

Antibiotic penetration into the CSF is generally thought to be increased when there is a breakdown in the blood-brain barrier due to meningeal inflammation in persons with TBM.^17^ We therefore sought to mine concomitantly collected high-resolution metabolomics and cytokine data to determine whether any soluble immune mediators were associated with CSF drug concentrations. We used mixed effects linear models to determine which cytokines and metabolites with known chemical identities were most strongly associated with CSF concentrations of each antibiotic. In **Figure 5**, we show the metabolites and cytokines significantly associated with the concentration of at least one antibiotic using a false discovery correction of q<0.05. We found that carnitines were significantly associated with CSF concentrations of ethambutol and imipenem at q<0.05, as well as isoniazid, linezolid, pyrazinamide, and rifampin at an unadjusted p<0.05. However, the directionality of the association was inconsistent. While increased CSF concentrations of carnitines were associated with increased concentrations of ethambutol and imipenem, they were associated with lower CSF concentrations of isoniazid, linezolid, pyrazinamide, and rifampin. This suggests that different metabolic states in the CNS may be associated with increased drug penetration for certain classes of antibiotics while decreasing CSF drug concentrations for others.

**Figure 5 –.**
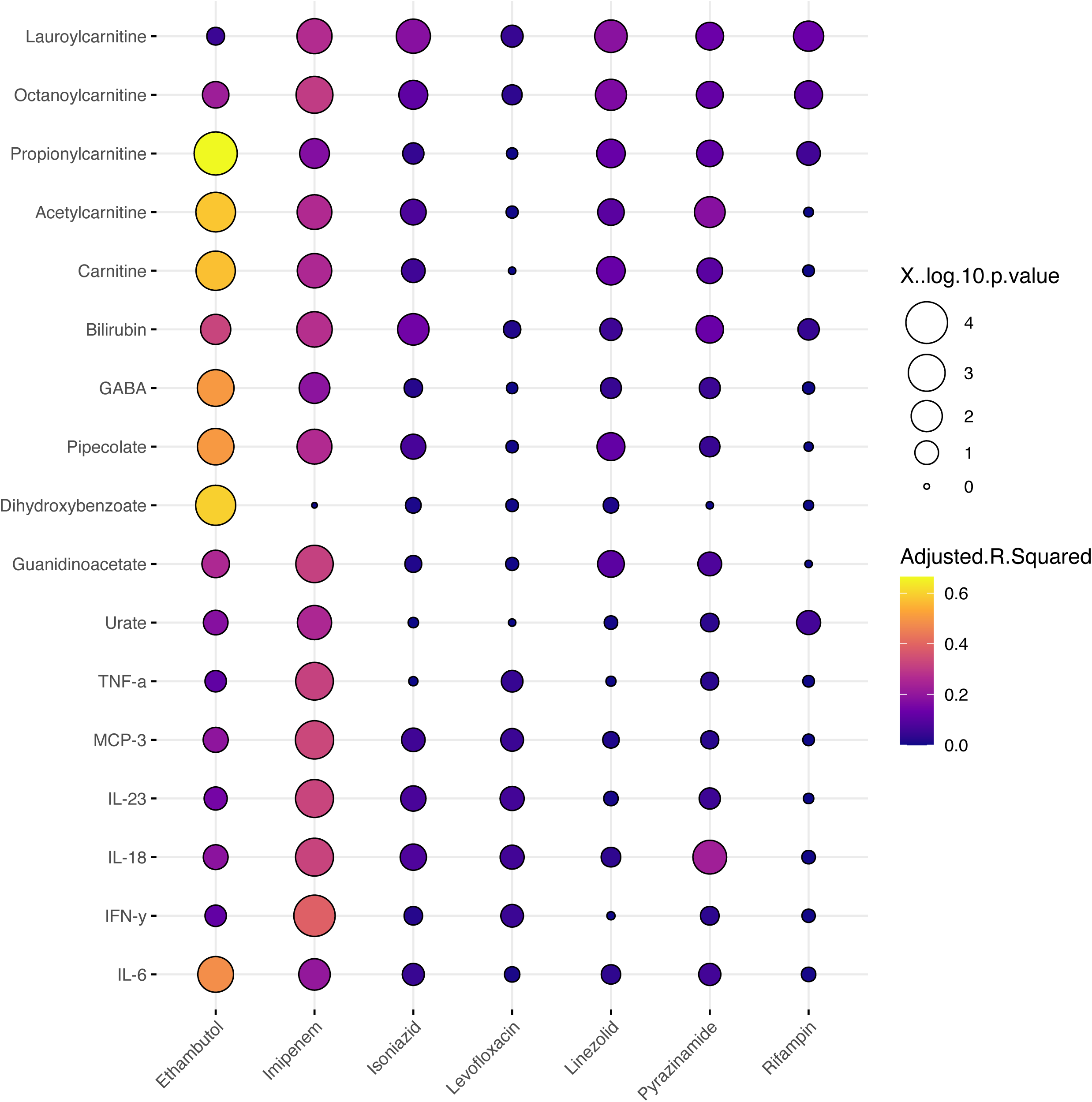
The bubble plots represent the adjusted R^2^ values measuring the association between anti-TB drug concentrations and soluble immune mediators (metabolites, cytokines, and chemokines) in the cerebrospinal fluid. The color scale for each dot represents the adjusted R^2^ value and dot size represents the –log P value for each association.

There were few significant associations between cytokine and antibiotic concentrations in the CNS. Increased concentrations of pro-inflammatory cytokines and chemokines including TNF-a, MCP-3, IL-6, IL-18, and IFN-y were associated with increased concentrations of imipenem, but otherwise had little relationship to CSF antibiotic concentrations. The only exception was IL-6, which was significantly associated with CSF ethambutol concentrations. These data indicate that an inflammatory CSF milieu had minimal impact on CSF concentrations anti-TB antibiotics.

## DISCUSSION

In this TBM pharmacometabolomics study, we show there is significant interindividual variability in CSF concentrations of anti-TB antibiotics. We further show that differences in serum drug concentrations account for much of the variability in CSF concentrations of linezolid, isoniazid, and cycloserine, but have a less strong relationship with CSF concentrations of imipenem, rifampin, moxifloxacin, and ethambutol. Finally, we demonstrate that CSF drug concentrations may be affected by the CSF metabolic milieu and have little association with concentrations of proinflammatory cytokines and chemokines. Together, these data show that a complex mix of factors contribute to CSF concentrations of anti-TB antibiotics. For many antibiotics, factors such as CNS inflammation and serum concentrations, which are traditionally thought to be primary drivers of drug concentrations in the CNS, may play only a minor role.

While serum concentrations of anti-TB drugs are known to vary widely from person to person,^16^ we show equally wide variability in CSF drug concentrations in those treated for TBM. Across the study population we found greater than 2-fold differences in CSF concentrations of all anti-TB drugs studied. This was true both two and six hours after the dose of each drug. Such variability in the CSF concentrations of anti-TB drugs has been shown in other studies, most notably among 237 patients (>700 CSF samples) included in a pharmacology sub study evaluating rifampin, isoniazid, and levofloxacin CSF concentrations.^18^ The substantial variation in CSF concentrations among study participants was evidenced by large 95% confidence intervals for both C_max_ and AUC parameters.^18^ Furthermore, interpatient variability in CSF rifampin^19^ and fluoroquinolone^20^ concentrations have been demonstrated in clinical trial dose finding and drug comparison studies, respectively. What has been less studied, is clinical or laboratory predictors of CSF drug concentrations. We found that serum concentrations are poorly predictive of CSF concentrations for many drugs including imipenem, rifampin, moxifloxacin, and ethambutol. Using the plasma rifampin AUC_0-24_ parameter, Dian et al, found a high correlation between plasma and CSF rifampin concentrations (Spearman’s ρ 0.7, p<.01).^19^ A better understanding of predictors of CSF drug penetration is needed to help both understand the underlying mechanisms of drugs reaching the site of disease and also to identify clinically useful markers to help guide optimal drug doing in TBM.

This study provides evidence that untargeted, high-resolution metabolomics can be an accurate way to capture drug concentrations of multiple anti-TB drugs including imipenem, ethambutol, isoniazid, linezolid, and pyrazinamide. This result is supported by studies showing untargeted metabolomics can also be an accurate way to capture exposure to xenobiotics in the environment.^8^ The ability to accurately quantify drug concentrations as part of an untargeted metabolomics platform is likely to create additional opportunities to understand drug concentrations and pharmacokinetics in the context of broader measurement of host metabolism. Understanding broad host metabolic responses to drugs, termed “pharmacometabolomics”, is an emerging area of study with great potential to improve our understanding of heterogenous host responses to medications including antibiotics and enhance personalized medicine.^11^ However, a limitation of the approach has been the need to separately perform targeted quantification of drugs and untargeted metabolomics. Our findings suggest that in many cases, important insights in pharmacometabolomics could be obtained from untargeted metabolomics data alone.

By simultaneously collecting pharmacology, metabolomics, and cytokine data in the present study, we were able to examine which soluble immune and metabolic mediators in CSF are associated with improved CNS penetration of TB drugs. The association between CSF concentrations of multiple antibiotics and carnitines suggests that a metabolic milieu with increased oxidative phosphorylation may impact antibiotic penetration in the CNS. Increased carnitines in the CSF, which transport fatty acids from the cytosol into mitochondria to be oxidized, may reflect improved cellular function and lower levels of glycolysis, which tends to produce greater inflammatory signaling in immune cells.^9,21,22^ Historically, greater inflammation in the CNS has been thought to be a catalyst for drug penetration across blood-brain and blood-CSF barriers.^17^ The present study found little relationship between CSF drug concentrations and pro-inflammatory cytokines. While elevated CSF concentrations of cytokines does not necessarily indicate increased meningeal inflammation, the findings do suggest that the relationship between CSF drug concentrations and inflammation is minimal. This is supported by prior work from our group, which indicated CSF concentrations of anti-TB drugs are roughly equal early in the course TBM treatment versus two or more months after treatment start, when meningeal inflammation would be expected to decrease.^5^ However, another explanation for these findings is that the CSF milieu remains highly inflammatory months after treatment start,^7^ which could mean CNS drug penetration is enhanced for an extended period of time.

This study is subject to several limitations. The small sample size of persons with TBM and low number of microbiologically confirmed cases from a single geographic region available for analysis in this study may limit generalizability. Further, the robustness of statistical inferences from such a small sample is limited. Though we demonstrate a strong association between CSF concentrations of multiple metabolites and concentrations of anti-TB drugs, the observational nature of the study precludes us from establishing a causal relationship. In future studies it will be important to evaluate the links between key metabolites and CSF drug penetration using animal models of TBM and larger, diverse human cohorts. Enhancing our understanding of how soluble mediators in the CNS impact drug penetration may lead to therapies that enhance drug delivery to this area or personalized dosing regimens for individual patients.

Overall, this study shows the interindividual variation in CSF drug concentrations among persons with TBM is high and potentially linked to the metabolic milieu of the CSF. Serum drug concentrations are only weakly associated with CSF concentrations for some drugs, and the amount of inflammation in the CSF appears to have only a minor role in enhancing drug penetration. Our results provide insight into the factors that impact CSF drug concentrations in TBM and indicate that improved understanding of the host metabolic response could provide targets to enhance drug delivery.

## MATERIAL AND METHODS

### Setting and Participants with TBM

Persons with TBM were enrolled from the National Center for Tuberculosis and Lung Diseases (NCTLD) in Tbilisi, Georgia as part of a clinical pharmacology study evaluating the penetration of anti-TB drugs into the CNS.^5^ Patients aged ≥16 years treated in the NCTLD adult TBM ward from January 2018 to December 2019 were eligible for inclusion. All patients suspected of having TBM underwent a lumbar puncture; acid-fast bacilli (AFB) staining, liquid and solid culture, and Xpert MTB/RIF assay were performed on CSF. As per standard of care for patients hospitalized with TBM at NCTLD, lumbar punctures were performed at approximately 7, 14, and 28 days after TBM treatment initiation and monthly for as long as patients were hospitalized to follow CSF cell and protein counts in response to treatment.^5,14^ Treatment regimens were selected by treating clinicians based on treatment history, comorbidities, and drug susceptibility results when available.^14^ All patients also received a 6–8-week course of dexamethasone (400-1200 mg). Written informed consent was obtained from all study participants, and study approval was obtained from the institutional review boards of Emory University and the NCTLD.

### Sample Collection and Drug Quantification

Blood samples were collected at 2 and 6 hours after drug administration at each timepoint. For CSF collection, 3 mL was collected at each time point, alternating between 2 and 6 hours after drug administration to capture early and delayed drug penetration into the CSF. All centrifuged blood as well as CSF samples were stored at – 80°C at the NCTLD until shipped to the Infectious Diseases Pharmacokinetic Laboratory at the University of Florida, where drug concentrations were quantified. Total concentrations of each drug were measured using validated liquid chromatography tandem mass spectrometry assays. The assays were validated for human plasma, and cross checked for matrix effects using artificial CSF. The analyses were performed on Thermo Scientific TSQ Endura or TSQ Quantum Ultra mass spectrometers.

### Metabolomics analysis

De-identified CSF samples were randomized by a computer-generated list into blocks of 40 samples prior to transfer to the analytical laboratory where personnel were blinded to clinical and demographic data. Thawed CSF (65 μL) was treated with 130 μl acetonitrile (2:1, v/v) containing an internal isotopic standard mixture (3.5 μL/sample), as previously described.^23^ Samples were centrifuged and supernatants were analyzed using an Orbitrap Q Exactive Mass Spectrometer (Thermo Scientific, San Jose, CA, USA) with dual HILIC positive and c18 negative liquid chromatography (Higgins Analytical, Targa, Mountain View, CA, USA, 2.1 x 10 cm) with a formic acid/acetonitrile gradient. The high-resolution mass spectrometer was operated over a scan range of 85 to 1275 mass/charge (*m/z*).^24^ Data were extracted and aligned using apLCMS ^25^ and xMSanalyzer ^26^ with each feature defined by specific *m/z* value, retention time, and integrated ion intensity.^24^ Three technical replicates were performed for each CSF sample and intensity values were median summarized.

### Cytokine detection

The commercially available U-PLEX assay by Meso Scale Discovery (MSD) was used for CSF cytokine detection. This assay allows for the evaluation of multiplexed biomarkers by using custom made U-PLEX sandwich antibodies with a SULFO-TAG conjugated antibody and electrochemiluminescence (ECL) detection. Direct quantitation of cytokines was performed using standard curves generated by 4-fold serial dilutions of standard calibrators provided by MSD. Plates were read on the QuickPlex SQ 120 using Methodical Mind^TM^ software and plate data analyzed using Discovery Workbench^TM^ software (MSD). ^27,28^

## Statistical Analysis

Descriptive statistics were provided for baseline characteristics, and the concentrations of anti-TB antibiotics in patients’ CSF were summarized for each drug at both 2-hour and 6-hour time points following the most recent antibiotic dose. We applied a linear mixed regression model, adjusting for sampling time (2 or 6 hours),^27^ and reported the adjusted R-squared values of each pairwise comparison of models, whether they included or excluded serum concentration^29^. Similarly, to determine associations between CSF drug concentrations and metabolite and cytokine concentrations, we applied a linear mixed regression model, adjusting for sampling time (2 or 6 hours), and reported the adjusted R-squared values of each pairwise comparison of models, whether they included or excluded the concentration of a particular metabolite or cytokine^29^. A false discovery rate (FDR) correction was employed to account for multiple comparisons when examining associations between anti-TB drugs and metabolites and cytokines.^28^ All analyses were conducted using R version 4.2.1.

## Data Availability

All data produced in the present study are available upon reasonable request to the authors.

## Acknowledgments

The authors thank the physicians, nurses, and staff at the NCTLD in Tbilisi, Georgia, who provided care for the patients with TBM included in this study. Additionally, the authors are thankful for study participants with tuberculosis meningitis who were willing to participate in the study and help contribute meaningful data that may help future patients with the same illness.

